# Understanding the impact of the COVID-19 pandemic and its control measures on women and children: A Zimbabwe case study

**DOI:** 10.1101/2024.06.03.24308400

**Authors:** Tinotenda Taruvinga, Rudo S. Chingono, Ioana D. Olaru, Kenneth Masiye, Claudius Madanhire, Sharon Munhenzva, Sibusisiwe Sibanda, Lyton Mafuva, Natasha O’Sullivan, Abdinasir Y. Osman, Kevin Deane, Tsitsi Brandson, Manes Munyanyi, Annamercy C. Makoni, Solwayo Ngwenya, Karen Webb, Theonevus T. Chinyanga, Rashida A. Ferrand, Justin Dixon, Katharina Kranzer, David McCoy

## Abstract

COVID-19 presented countries with unprecedented health policy challenges. For low-income countries in particular, policymakers had to contend with both the direct threats posed by COVID-19 as well as the social, educational, and economic harms associated with lockdown and other infection prevention and control measures. We present a holistic and contextualised case study of the direct and indirect impacts of COVID-19 on women and children, with some assessment of their uneven distribution across socio-economic, age and gender groups. We used different types of primary and secondary data from multiple sources to produce a holistic descriptive analysis. Primary data included: qualitative data obtained from 28 in-depth interviews of key informants, six focus group discussions; and 40 household interviews. We also extracted data from government reports and announcements, the District Health Information Software version 2 (DHIS2), newspaper articles and social media, as well as from published research articles. Our findings show that the direct and indirect adverse impacts of COVID-19 were compounded by many years of severe political economic challenges, and consequent deterioration of the healthcare system. The indirect effects of the pandemic had the most severe impacts on the poorest segment of society and widened age and gender inequalities. The pandemic and its accompanying infection prevention and control measures negatively affected health service delivery and uptake. The management of COVID-19 presented enormous challenges to policymakers and public health specialists. These included managing the greatest tension between direct and indirect harms; short-term and long-term effects; and the unequal distribution of harms across different segments of society.

## INTRODUCTION

The SARS-CoV-2 (COVID-19) pandemic presented all countries with major health challenges [1,2]. These included ensuring adequate epidemiological surveillance of COVID-19, implementing infection prevention and control (IPC) measures, providing care for both COVID-19 and non-COVID-19 patients, and procuring including delivering COVID-19 vaccines. Low- and-middle-income countries (LMICs) were also faced with having to implement draconian lockdown measures without having the resources to mitigate their social, educational, and economic harms [3,4], and the disruption caused to normal healthcare provision [5,6].

Importantly, countries had to consider how the harms associated with the pandemic would be unevenly distributed [1,7,8]. For example, wealthier households would be better able to withstand the psychological and emotional stresses of lockdown compared to poorer households [6,9]. Additionally, there were reasons to think that women and girls would experience lockdown measures differently from men and boys [8]. Finally, the early observation that infection fatality rates varied substantially across age groups raised challenges to policymakers about balancing the different needs of children and adults [10,11].

We conducted research aimed at presenting a holistic and contextualised case study of the direct and indirect impacts of COVID-19 on women and children in Zimbabwe, with some assessment of their uneven distribution across socio-economic, age and gender groups. As is typical of case studies seeking to understand complex phenomena, we used different types of primary and secondary data from multiple sources [12] to produce an integrated descriptive analysis of the COVID-19 epidemic, the policy responses to it, and their impacts on maternal, child and women’s health.

## METHODS

To produce a chronological narrative account of policies related to COVID-19 from February 2020 to August 2021, we collected data from government policy documents and announcements, newspaper articles, social media, and the Oxford COVID-19 response tracker (OxCGRT), a project that collated data on COVID-19 control measures from countries across the world [13].

To present an account of the COVID-19 pandemic, we obtained official data on the number of COVID-19 tests done, confirmed cases and deaths from the daily situational reports and the official Twitter (now X) handle of the Ministry of Health and Child Care (MoHCC). Given the limited disease surveillance and low testing rates, we also drew on data from other research to assess COVID-19 transmission in Zimbabwe.

To describe the effects and impacts of COVID-19 and its control measures on maternal and child healthcare, we extracted routinely collected health service data on antenatal care (ANC), growth monitoring of children and childhood vaccinations from the District Health Information Software version 2 (DHIS2) for the period January 2016 to August 2021 in Bulawayo and Harare metropolitan provinces. Harare the capital city and Bulawayo the second biggest city in the country have estimated populations of 1,896,134 and 676,650, respectively, and together accounted for about 30% of the country’s recorded COVID-19 cases [14]. Primary health services in the two cities are primarily administered by the city council whilst tertiary and quaternary hospitals are administered by the central government. We selected 20 public primary health clinics in Harare and 18 in Bulawayo to study trends in healthcare provision and utilisation.

We selected two child and two maternal services: growth monitoring (GM), child vaccinations, antenatal care (ANC) and HIV testing of pregnant women. Several data items are routinely collected to monitor child vaccination services indicators, and we chose to analyse trends in the number of children aged 9-12 months who have received their final measles, mumps, rubella (MMR) vaccine which is captured in the DHIS2 as ‘primary course complete’ (PCC). For maternal health services, we chose to analyse trends in the number of pregnant women attending a 4^th^ ANC visit (a proxy indicator of adequate ANC visits according to the WHO) as well as the number of women who received an HIV test at their first ANC visit. However, data for the latter was only available from 2019 onwards.

Changes over time in the four indicators of healthcare utilisation are shown graphically for each clinic, stratified by city, with trend lines using loess smoothing and 95% confidence intervals. Clinic-level changes in healthcare utilisation are shown as ratios computed by dividing monthly counts by the clinic-specific monthly average for the period starting January 2016 up to the end of 2019. The ratios were colour-coded using shades of orange to mark counts that were lower than the pre-COVID-19 average and shades of green for higher counts. Some clinics had missing values for some months, especially in 2021, and these were coloured in grey. These missing values include a combination of instances where there was no service uptake (e.g. the clinic was open but there was no service utilisation, or the clinic was shut) and when there was service uptake, but data were not recorded or uploaded onto the DHIS2.

We also collected primary qualitative data between April 2021 and December 2021 in Harare and Bulawayo. We conducted a total of 28 in-depth interviews (IDIs) of key informants drawn from community-based organisations (CBOs) (n=5), city health managers (n=5 Harare, n=3 Bulawayo), national programme managers and policymakers (n=5) and healthcare workers (n=3 Harare, n=7 Bulawayo). We further conducted two focus group discussions (FGDs) with healthcare workers from selected primary health facilities in Harare (n=12) and Bulawayo (n=11), and four FGDs of members of the public (two in Harare with 9 and 10 participants, and two in Bulawayo with 11 and 13 participants). In addition, we conducted 40 household interviews, evenly distributed across three economic strata (n=20 Harare and n=20 Bulawayo). Topic guides were used for all the qualitative interviews. Written informed consent was obtained from all the participants in the study.

Finally, we drew data from selected published literature to describe the historical and socio-economic context of the pandemic and add further information about the impact of COVID-19 on maternal and child health in Zimbabwe. We identified this literature from the database of PubMed and Google Scholar for papers using search terms ‘maternal and child health’, ‘COVID-19’ and ‘Africa’ or ‘Zimbabwe’ up to the period January 2023.

## FINDINGS

### 1. The context of Zimbabwe’s COVID-19 epidemic

Zimbabwe is a low-income landlocked country of about 15 million people, with 42% of the population under the age of 15 [14]. In 2019, its Human Development Index was only 0.571, ranking 150^th^ out of 189 countries [15]. Approximately 71% of the population lives below the international poverty line of $1.90 per day [16] leading to poor health indicators. Over 80% of income earners operate in the informal sector [17] which contributes to over 60% of gross domestic product (GDP) [18,19]. One in four of the urban population (about 1.25 million people) live in settlements with poor water and sanitation infrastructure as defined by UNICEF [20]. However, Zimbabwe has a highly literate population with 94% of men and women aged 15-49 years able to read, and 91% of all children of primary school age attending school [19]. Moreover, Zimbabwe’s Gender Development Index (GDI) was 0.952 in 2018, suggesting relatively good gender equity, with secondary school attendance among girls being higher than boys [21].

The country had experienced severe political and economic challenges for the three decades before the COVID-19 pandemic. An unsustainable external debt burden and a structural adjustment programme resulted in a severe contraction of public expenditure in the 1990s which weakened most public services [22]. The turn of the millennium saw the country enter a period of protracted political turmoil and the imposition of targeted sanctions by several European countries [22,23] further undermined the economy. Just before the pandemic, Zimbabwe’s GDP had declined by 11.3% in 2019 compared to 2018 and continued to contract in 2020 [24].

Although Zimbabwe’s health system was once considered among the best in Africa, [18,25] it deteriorated due to reductions in public spending, the out-migration of health professionals, mismanagement, and industrial action [26,27]. By 2021 the doctor and nurse persons ratios had deteriorated to 1:12,000 and 0.39:1,000 respectively, far below the World Health Organization’s (WHO) recommended developing countries’ minimum threshold density of 2.28 doctors, nurses and midwives per 1,000 persons [28]. Today, there are high levels of out-of-pocket payments and donor dependency [29]. The maternal mortality ratio was 458 per 100,000 live births [30] and the child mortality rate was 55 per 1000 live births [30] in 2020 [31,32].

On top of this chronic deterioration of the health system, there were a series of acute crises in the run-up to the pandemic (see Figure 1). This included doctors and nurses engaging in intermittent industrial action [33], typhoid and cholera outbreaks in 2018, a major natural disaster (cyclone Idai) [34] in 2019, an acute worsening of inflation from April 2020 causing shortages of fuel and water. The precarious state of the health system at the time of the pandemic was noted in our qualitative data.

> *“Most of our stream mates have gone to the UK where there are better jobs. Others are in the process of applying; we are all trying to leave for greener pastures (laughs). Those of us who stay, we are mostly working out of duty…people are disgruntled, and this can be sometimes seen through our attitudes and, umm, the strikes.”* (Nurse, HCW FGD, Bulawayo).

**Figure 1:**
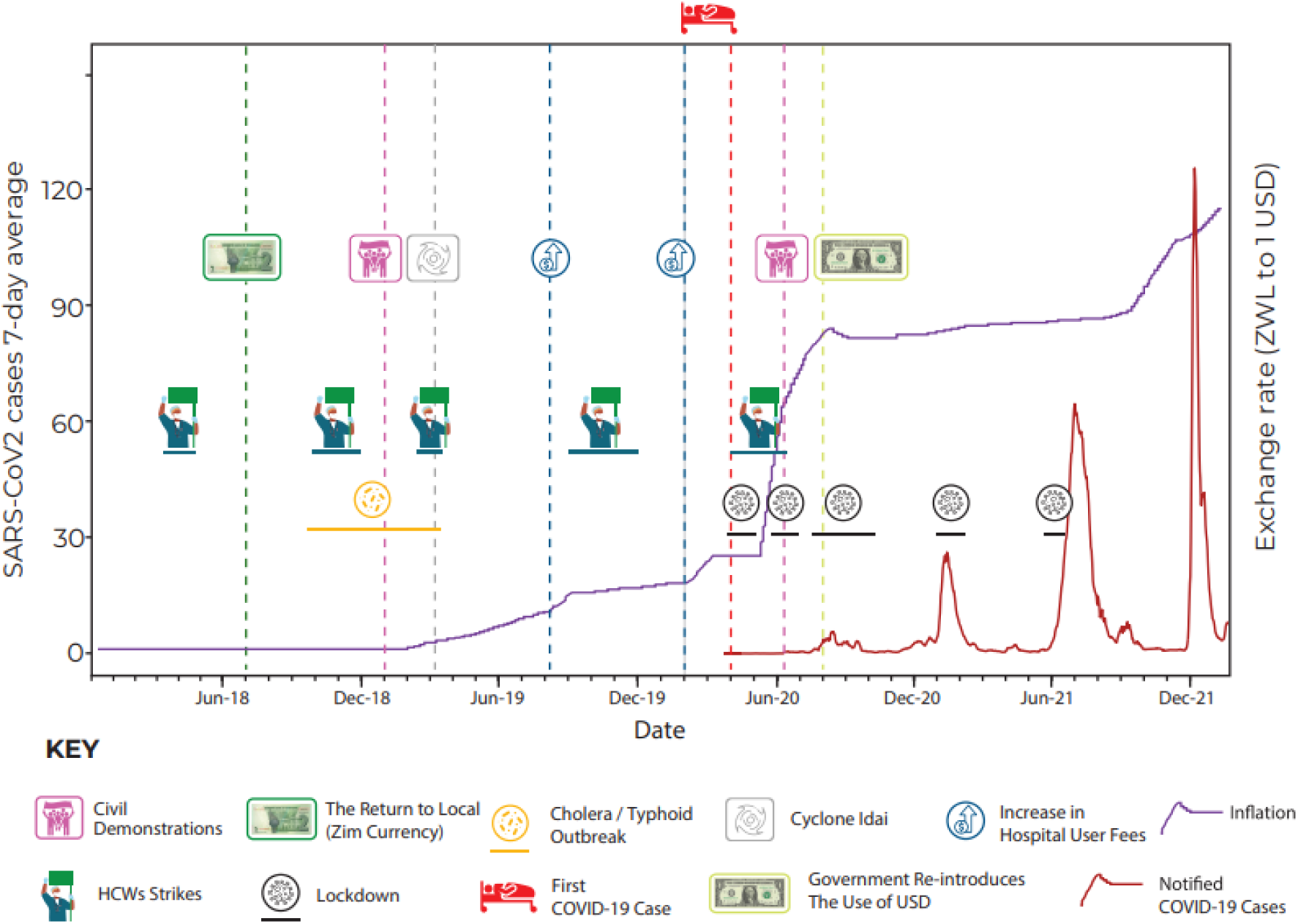
Timeline of key events concerning the period between January 2018 to December 2021.

> *"Unfortunately, in Bulawayo when COVID-19 was declared and it hit us, we were facing water challenges and other related things.”* (Doctor, KII, Bulawayo).

### 2. Evolution of the COVID-19 epidemic and control measures

#### (i) January to May 2020

Following WHO’s call to prepare for the containment of COVID-19 in January 2020, the government of Zimbabwe quickly started developing a National Preparedness and Response (NPR) Plan and established an Interagency Coordination on Health and Epidemic Preparedness and Response Task Force [35]. Screening for SARS-CoV-2 infections at major entry points was quickly implemented for all incoming individuals. A series of inter-ministerial meetings on COVID-19 also took place in February and March with the involvement of the private sector, civil society, academia, professional associations, technical and donor partners and the media.

The NPR Plan was informed by a Preparedness and Response Plan for Pandemic Influenza A H1N1 formulated in 2009, recent simulation exercises, a comprehensive review of the 2018 cholera outbreak and a rapid assessment of the state of preparedness at major points of entry into the country and major health facilities in February. The Plan was aligned with WHO’s COVID-19 Strategic Preparedness and Response Plan and finalised in March.

On March 17^th^, a state of national emergency was declared prior to any detectable surge in SARS-CoV-2 infections. In fact, the first case of COVID-19 was only reported on 20^th^ March in an individual who had recently returned from the United Kingdom. Two days later, two additional cases, imported from the United States, were documented. The first recorded fatality occurred on 23^rd^ March [36], the same day the Civil Protection (Declaration of State of Disaster: Rural and Urban Areas of Zimbabwe) (COVID-19) Notice was issued [37].

A Public Health Act (COVID-19 Containment and Treatment) was also enacted on the 24^th^ of March, mandating compulsory testing of individuals suspected of infection and the quarantine of those who tested positive [38]. Subsequently, a Statutory Instrument (SI) was introduced imposing a 21-day lockdown from the 28^th^ of March during which time public gatherings of more than two people were prohibited. While ‘essential services’ were allowed to operate, retail outlets were only allowed to open during specific hours, with mandatory screening and hand sanitising at entrances. Although initially, schools and universities were slated to remain open until the end of the term, a later announcement mandated the closure of all educational institutions from 24^th^ March 2020. The sale of alcohol was banned, and the stockpiling of medical supplies and food was explicitly prohibited with strict penalties for any violations.

During this phase, the government also designated certain health facilities as quarantine and isolation centres, adopted and adapted WHO’s COVID-19 case management guidelines [39], organised training for health workers and upgraded several government and private hospitals. Surveillance teams were created to facilitate case investigation and contact tracing. Over 4,000 health sector posts were unfrozen, and an additional 200 new medical posts were created and funded through a reallocation of funds [40] from other departments and from the proceeds of the 2% Intermediated Money Transfer Tax which is normally used for social protection and capital development projects. However, despite efforts to strengthen the health system’s capacity to respond to COVID-19, the shortage of PPE and inadequate pay led many healthcare workers to feel vulnerable and undervalued [41] and precipitated a fresh round of industrial action beginning on 25^th^ March with a nurses’ strike [42].

> *“An association representing the City of Harare nurses went around to check if there was adequate PPE…After they observed that there was nothing that’s when they called for the nursing staff or for the healthcare worker to stop going to work, tools down guys there is no PPE until proper adequate PPE is in place. So, you find that most nurses of the city of Harare went home and stayed…The situation was also aggravated by the fact that we were being poorly remunerated, we were not getting our salaries on time.”* (Nurse, HCW FGD, Harare).

Lockdown measures were eventually extended for a further four weeks to 17^th^ May, although from 3^rd^ May, the formal commercial and industrial sectors were allowed to operate between 08:00 and 15:00, provided IPC measures were in place. Throughout this period, numerous SIs were gazetted to amend or extend the initial lockdown measures. For example, on April 3rd, facemasks were made compulsory in public and transport services were required to check the temperatures of boarding passengers and implement disinfection protocols on all vehicles. While initially, residents were allowed to return home from abroad provided they quarantined at designated facilities, on May 4^th^ all international travel was banned. Land border crossings at Beitbridge (bordering South Africa) and Plumtree (bordering Botswana) were also closed, and internal travel by road was restricted, with exemption letters required at road checks.

Throughout this period, reported case numbers remained low. When lockdown was lifted on May 17^th^, Zimbabwe had only recorded 46 cases and six deaths. However, because public sector testing capacity was limited and the cost of private testing was unaffordable for most people, the true number of cases during this period is unknown. Further, there were no formal or informal reports of any surge in hospital admissions and mortality during the period.

#### (ii) June 2020 – November 2020

After the first lockdown was lifted, the number of recorded cases remained low for about eight weeks. Despite this, the government imposed a two-week lockdown from June 20 to July 4. This followed extensive civil society protests following the arrest of the health minister on allegations of graft. It is widely believed that the lockdown was used to suppress political opposition rather than the epidemic [43,44], with many protestors arrested for contravening the new COVID-19 regulations.

However, when case numbers started rising, the government introduced a third lockdown on July 24^th^. Night-time curfews (from 6 pm to 6 am) and travel restrictions were implemented. However, essential services continued to operate as usual and non-essential businesses and low-risk sports were allowed to operate from 9 am to 3 pm. Restaurants, hotels and other tourism services were allowed to operate at 50% capacity; and schools were kept open until July 30^th^ to allow the completion of national examinations. Although the rise in case numbers was partly due to increased testing capacity [45] there was some evidence of a real increase in the incidence of infection with rising COVID-19 mortality and hospitalisations [46] including some high-profile fatalities.

The third lockdown continued until September but with a progressive easing of restrictions as case numbers started falling. Government quarantine centres were phased out in August and replaced with self-isolation in private premises, and a phased opening of schools commenced on the 8^th^ of September and was completed in November. The sale of alcohol for consumption off-premises was allowed on 30 September, and international airports started operating again in October. While initially, only Zimbabwean residents were allowed to return (provided they tested negative and self-quarantined at home for 14 days), entry was later allowed for foreign travellers. In December land ports of entry were opened.

#### (iii) December 2020 – April 2021

In late November 2020, a new wave of cases emerged, driven by the Beta variant [47]. This coincided with the reopening of schools, where some large outbreaks were reported [48]. As numbers rose, and with several high-profile deaths including that of five cabinet ministers [49], the government introduced a fourth national lockdown for 30-days on January 3, which was extended to February 16 and then again to February 28. The government closed schools, prohibited gatherings, reintroduced curfew hours and stopped intercity travel. Mask-wearing, hand sanitisation/washing, and temperature checks remained mandatory in public. As the country started recording a reduced number of cases, lockdown measures were eased in March, inter-city travel resumed, and schools re-opened in a phased manner. April saw a relaxation of most remaining IPC measures although international travel to and from countries such as India that had reported the Delta variant were not permitted while travellers from other countries were required to present a negative COVID-19 test result and self-quarantine at home or in designated quarantine centres for 10 days [50].

During this period the country also began a COVID-19 vaccination program with a target of vaccinating 10 million people by December 2021 [51]. The first phase prioritised frontline healthcare workers, the elderly, and those with chronic medical conditions [52]. The second phase which began in March 2021 extended coverage to uniformed forces, all civil servants and those offering essential services in the private sector. The last phase was introduced in July and extended coverage initially to everyone over 18 years, then to anyone over 16 years and finally to anyone over 12 years [53].

#### (iv) May 2021 – September 2021: Third wave and fourth lockdown

In the middle of May, the new (Delta) variant was detected in the Kwekwe district, Midlands province. A new SI was introduced to allow for a localised lockdown of the Kwekwe district and then in two (Hurungwe and Kariba) other districts [54]. Eventually on June 27th, due to rising cases, a fifth full lockdown was implemented across the whole country. However, despite the increase in cases, the government came under pressure from parents and opened schools in the first week of August 2021. This was followed on August 10 by the reopening of some social activities with, for example, vaccinated congregants being allowed to attend church services.

Meanwhile, by the end of September, 3,051,371 first vaccine doses and 2,211,880 second vaccine doses had been administered nationally translating to national coverage of 35.7% and 25.8% respectively [55]. At about the same time the Global Fund pledged USD 75-150 million [56] to be used to mitigate the impact of the COVID-19 pandemic on HIV, tuberculosis, and malaria. From September onwards, the country gradually moved into a post-pandemic phase even though there was no official declaration of the pandemic being over. However, the government emphasised more on ‘living with the new normal’.

### 3. Policy implementation

According to our key informants, a notable feature of the response to COVID-19 was that policies were made and implemented in a centralised and top-down manner with little bottom-up and contextualised input.

> *“There was centralisation of decision making and policy formulation …. everything was moved to the centre …and ours was just to implement. Then we had a situation where there was a take-over … of local authority institutions by central government. It had its downside and upside. But the formulation of that policy, unfortunately, we didn’t input in."* (Doctor, KII, Bulawayo).

> *“Let’s not take that one size fits all or… maybe some African countries are doing this, you just follow suit and do this …. We should design our measures to suit our people, to suit our needs and to suit our country and our resources.”* (Nurse in charge, IDI, Harare).

> *“I still insist that we must not have a lockdown that stretches from Zambezi to Limpopo, that is being the same. Let’s modify it and say we are now in Gweru. How do we make this lockdown work? When we get to Chirundu, there it must not look like the one in Chiredzi. There must be that distinction. I feel that with the localised input and the modifications we make, the lockdowns will be more effective. Such that it will allow us to even open the economy in one part of the country whilst the other part is under lockdown.”* (Doctor, KII, Bulawayo).

Informants also noted that public health communication (mainly conducted through the Zimbabwean broadcasting cooperation’s television and radio channels) was not fully effective and that misinformation and disinformation on various social media platforms contributed to poor compliance with COVID-19 control measures, despite rapid response teams being established to respond swiftly to rumours and false information, albeit limited capacities to deal with all the rumours due to shortage of human and material resources. Others indicated that many people felt overloaded with conflicting information [41].

> *“What was our challenge was people didn’t have information. So, you would just see like a uniformed person come to you to hit you and you don’t have information, especially at the beginning. People were just being hit for COVID, but if you would have educated people … “this is COVID, people are affected this way, we need to prevent using these methods” … I think we are an educated nation that can actually follow through and take instructions”* (CBO representative, KII, Bulawayo).

> *“We communicated, we talked, we tried to send messages, but I don’t think it was effective because people had their own expectations, and they had their own other sources of information. And they did not trust the figures that were coming out of the Ministry of Health and situational reports eeh however, localised or frequent, people still did not feel that they, they were in as much danger as it was being portrayed.”* (Doctor, KII, Bulawayo).

Another theme that emerged from our primary data was the impracticality and impossibility of implementing social distancing measures, especially in high-density and overcrowded informal settlements with inadequate water and sanitation infrastructure [57]. Other measures such as the use of hand sanitisers were unaffordable for many. According to one study in Harare and Mashonaland East, between 18% and 53% of healthcare workers reported a lack of soap, water, and masks across the period from June 2020 to the middle of 2021 [58]. Other studies similarly described how communities could not comply with social distancing measures because of the need to secure food, water, and income [41].

> *"We expect the government not to implement water cuts and to improve water supply during such a time because we are having a difficult time, social distance is not possible [in the queues], we are at risk of contracting COVID-19”* (Participant, household IDI, Harare).

> *“It was impossible to comply with the social distancing measure, at a time when there were food shortages and one had to stand in long queues. I still remember there was a time I went to a queue to get mealie-meal at Malbereign. The soldiers had to come and space out the people.”* (Participant, household IDI, Harare).

Another cause for poor compliance with lockdown measures was the inability to monitor and enforce travel restrictions. Despite the police and army being deployed, resources were insufficient to enforce quarantine and ensure adherence to travel restrictions. For example, despite major points of entry being manned by armed security, many people were able to enter the country illegally without being quarantined [59].

The relative ineffectiveness of lockdown measures may also be deduced from seroprevalence studies which found that by the end of March 2021, a high proportion of the population had been infected. In one study of randomly selected households from three high-density communities in Harare, seroprevalence was found to have risen from 19% in November-December 2020 to 53% in Feb-April 2021 [59], suggesting high levels of community transmission during the second wave. A survey of healthcare workers as early as June 2020 reported that 39.1% had a history of COVID-19 symptoms and a SARS-CoV-2 seroprevalence of 8.9% [60].

A systematic review of seroprevalence studies in Africa between January 2020 and December 2021 also estimated that seroprevalence in Southern Africa was 56.1% (95% CI 44.6% - 66.9%) by the third quarter of 2021, with lower rates in rural areas and high heterogeneity between countries [61]. Of note is a study from Malawi that found seroprevalence increasing from 11% in December 2020 to 65% in April 2021 [61]. A similar increase in seroprevalence was observed in a study from Harare conducted between November 2020 and April 2021 [59]. The reported data also indicated a less severe disease profile in Africa with more asymptomatic cases compared to other parts of the world [62].

Another feature of the policy response to COVID-19 was the government’s efforts to mitigate the social and economic harms of lockdown. Crucially, in May 2020 a COVID-19 Economic Recovery and Stimulus Package worth about 9% of GDP was announced and included income support for all vulnerable groups (US$5 monthly for everyone in a vulnerable household) and fiscal support to key sectors of the economy such as manufacturing, agriculture, mining, and tourism [63]. The government also put in temporary measures to defer rent and mortgage payments during lockdown. However, the income support payments were not fully implemented or implemented for long enough. Dzawanda citing Buckle and Mpofu reported that the government’s promise of a once-off allowance for vulnerable people was only partially delivered and that four months into the lockdown, only 202,000 of the 1 million eligible households had received the allowance [18]. Similarly, a promised compensation scheme for civil servants who lost their lives while on duty was only partially delivered, although all civil servants were given a US$75 COVID-19 monthly allowance for over 36 months in addition to their salaries.

> *"I think maybe what probably might have lacked…are the necessary support systems to make sure that we take care of the most vulnerable in society, for example, if we say there is no public transport that is going to be operating, we should then be able to provide transport for those that want to seek health services.”* (NGO representative, KII, Harare).

> *“For an economy that is largely informal, if we then say we are closing everything else all the markets non-informal business and everything else, we should then put in place a system where we can then support those in the informal sector so that at the end of the day, they don’t uhm starve.”* (CBO representative, KII, Bulawayo).

### 4. Impact of COVID-19 control measures

This section looks at the pandemic’s impact on maternal and child health (MCH) services provision and utilisation; interpersonal and household relations; social and economic well- being; and child education and well-being.

#### (i) Maternal and child healthcare

Our analysis of routinely collected data from July 2016 to July 2021 in Harare and Bulawayo found evidence of a reduction in healthcare utilisation. Figure 2 presents data on the number of fourth antenatal care visits by pregnant women and the number of HIV tests conducted among pregnant women at their first antenatal visit.

**Figure 2:**
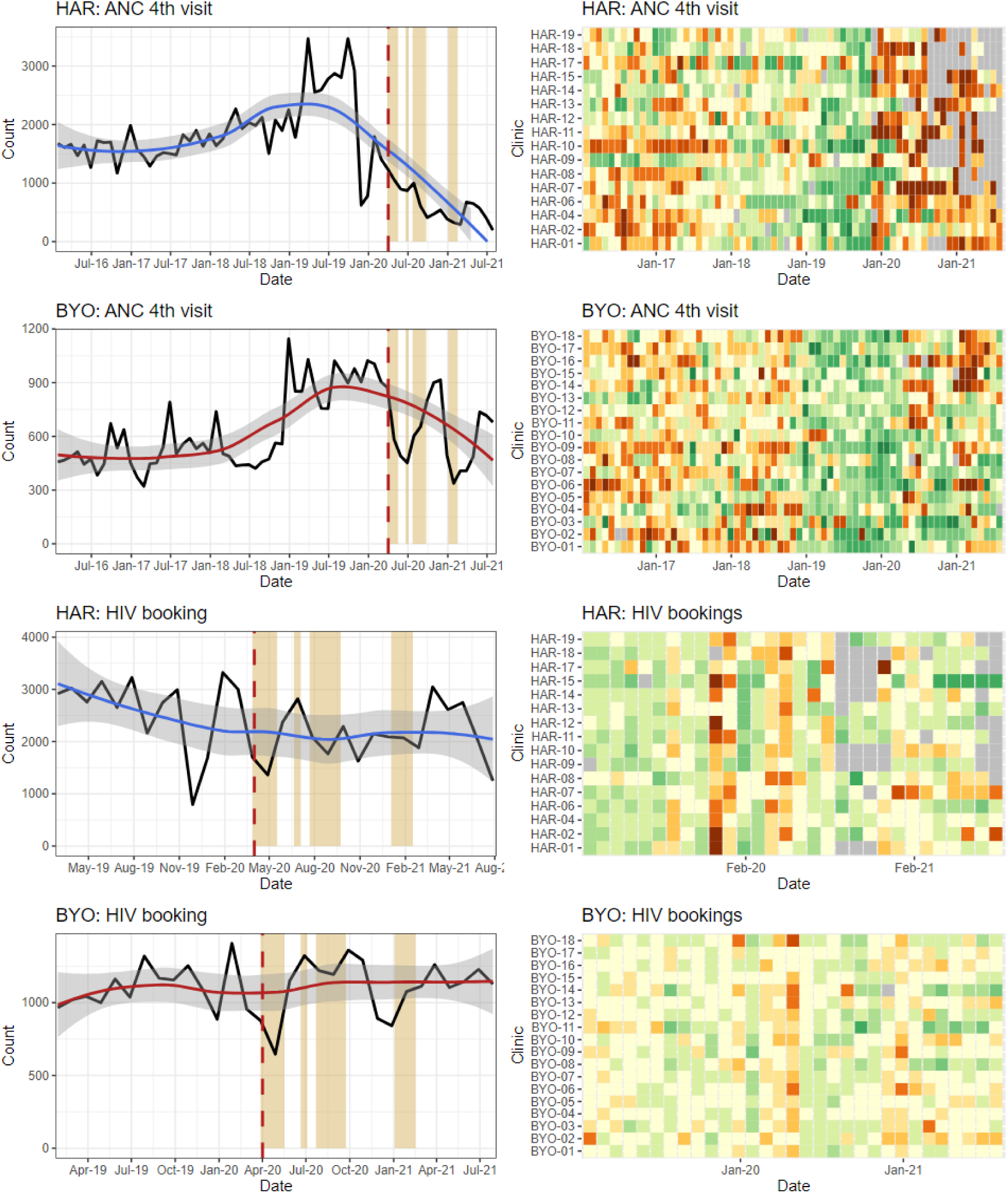
Source: Zimbabwe District Health Information Software Version 2 maternal indicators.

The data show a reduction in the number of ANC fourth visits in both provinces, with a steeper decline in Harare compared to Bulawayo. However, there was no observable reduction in the number of HIV tests on women attending antenatal care. Key informants had explained that the HIV programme which is largely donor-funded andrelatively well resourced was better able to maintain their pre-COVID-19 level of service. In one study conducted in Harare, the number of pregnant women accessing HIV services was shown to have stayed the same except for a slight decline during the first lockdown [5].

Figure 3 presents data on the number of GM visits by children under-5 years and the number of child PCCs. The data show that the number of GM visits decreased in both Harare and Bulawayo after the first COVID-19 case, and that in between lockdowns, GM visits rebounded to some extent in Bulawayo but not in Harare. The number of PCCs decreased substantially in Harare from March 2020 onwards but not in Bulawayo.

**Figure 3:**
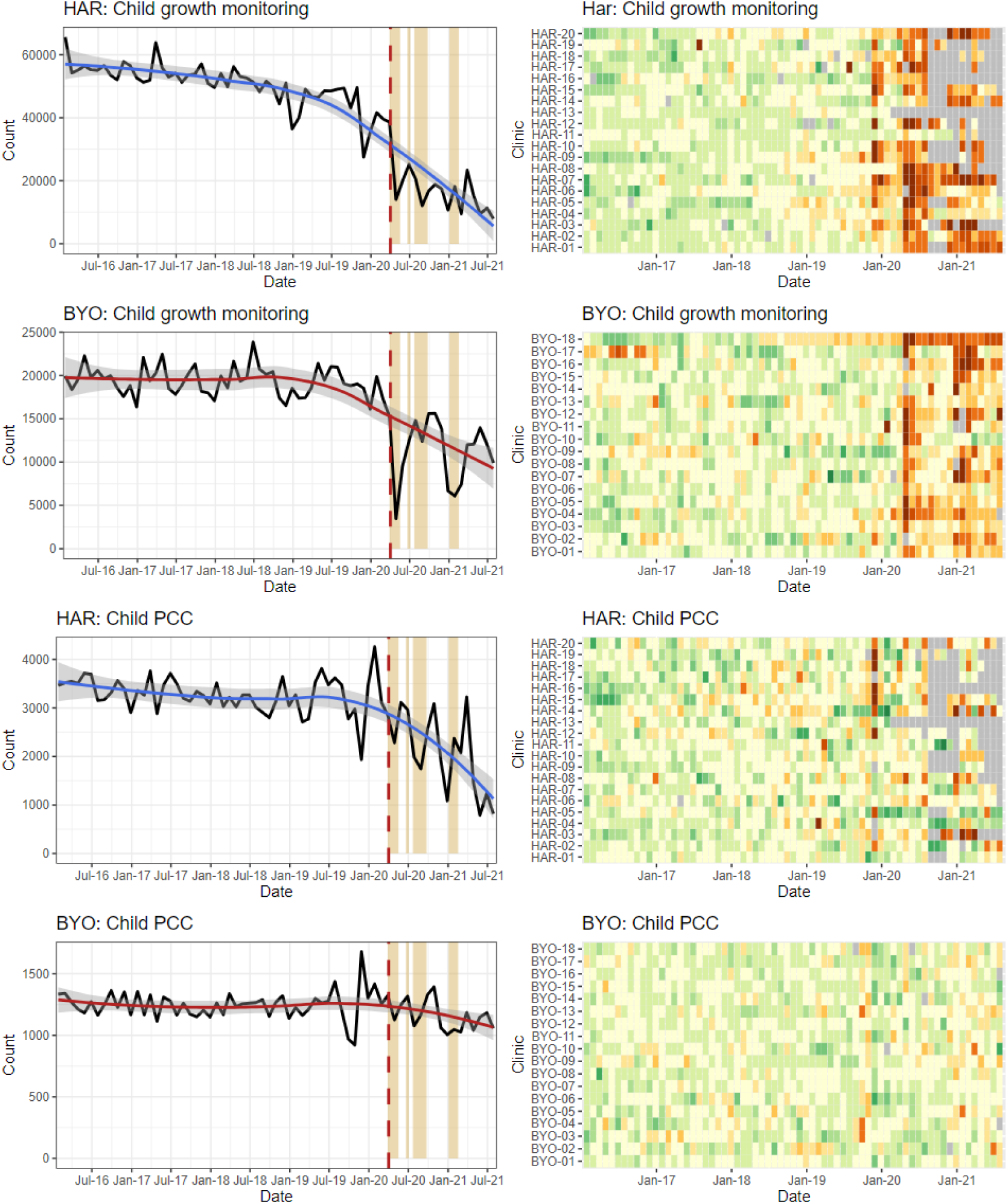
Source: Zimbabwe District Health Information Software Version 2 child indicators data.

Other studies also report that healthcare utilisation declined in 2020 and 2021 compared to earlier years [64,65]. One study from Harare found that the number of people registered for TB treatment had decreased by 34% and the number of individuals tested for HIV had dropped by 63% in 2020 and 2021 compared to the period between 2016 and 2019 [66]. TB treatment completion was found to have decreased from 81% to 70%, while treatment of people living with HIV with antiretroviral therapy dropped from 96% to 92% [66,67]. In a study at Mpilo Hospital in Bulawayo, although the mean number of monthly deliveries reduced from 747 in the first quarter of 2020 to 681 in the second quarter, the overall reduction was not substantial [68].

Our qualitative data pointed to several reasons for the reduction in healthcare utilisation. In some places, clinics were closed because staff went off sick because of COVID-19 [69], or because of industrial action. Some informants explained that healthcare utilisation also dropped because health facilities were perceived as “hotspots” for COVID-19 transmission, especially those that had been repurposed for COVID-19 services.

> *“People are not coming as they used to because staff tested positive and the community heard about it, and so maybe they are fearing that maybe when they come here, they might eh contract COVID-19"* (Nurse, IDI, Harare).

> *“I have a young sister who was pregnant at the time… she had to go to Murehwa because we thought it was safer than here in Harare, …"* (Participant, Household IDI, Harare).

Police roadblocks and checkpoints were highlighted as barriers to healthcare due to fear and unwillingness to disclose one’s health status. Healthcare workers reported finding it difficult to access public transport because they were perceived to be a risk to others, especially whilst wearing a uniform.

> *“You find someone could not access the hospital easily especially someone from the rural areas … there was no transport for them to come. Even those who are in town they couldn’t access the hospital there was no transport… some mothers could not take their babies for review even to the baby clinics, though it was said that the baby card was also a pass for them.”* (Nurse, IDI, Harare).

> *“Then people started to have difficulties in attending health centres, funerals, no hospital visits, this is the period from July to August. Whereby we are saying even HIV status disclosure on a roadblock. People were now forced to disclose… So, this was a forced disclosure…”* (NGO representative, KII, Bulawayo).

> *"And other people ended up not even attempting to go …. we have other people who are on ARVs whom we know that have defaulted because they don’t want to be harassed because you gave them a lift and you get there, they are asked to produce a card by the police, at times they would want to hide their status and not produce that card so end up not going to the hospital.”* (Nurse, HCW FGD, Harare).

Health services were especially disrupted by operating times being reduced from 07:30-17:30 to 08:00-14:00 and by the capping of the number of patients seen per day. In some instances, operating hours were reduced even more because healthcare workers struggled to get to work or had to leave early to comply with the curfew.

> *“Yes, at the clinic, they would serve a certain number of people per day, so that people don’t get crowded there. So, if they reach the number of people that they wanted per day, some would be returned home and be advised to come the next day …”* (Participant, Household IDI, Harare).

> *“Our staff couldn’t get transport to work, no one was willing to give them a lift as people considered them to be a high-risk group…We also had 6 staff members who tested positive and the news spread in the local communities and people were avoiding this facility for fear of getting the virus…”* (Nurse in charge, IDI, Harare).

The pandemic diverted resources and attention away from MCH services. Informants reported that in some clinics, GM was completely suspended to allow for prioritisation of other health services. Stockouts particularly affected immunisation and family planning services [70].

> *“Then in those findings, it was noticed that there was growth monitoring reduction. Then we also go to maternity health services whereby during the COVID era, we are saying only ANC bookings were done. No subsequent visits were done, limited monitoring of BP checks. If you remember we were no longer doing these BP checks, no physical examinations were done. Then om HIV and AIDS testing, the numbers started to reduce, viral load collection we stopped doing it, CD4 count was stopped, the updating of green books was also stopped.”* (Nurse, IDI, Bulawayo).

Access to healthcare was also restricted by some facilities implementing a policy of patients, including pregnant women, showing a negative SARS-CoV-2 test before being admitted. This led to patients being turned away from hospitals and ambulances refusing to transport patients:

> *“Pregnant mothers were requested to have results for COVID-19 first before admission to a facility and the test price was very high some of the mothers could not afford it… and even the ambulance services would request you to produce results first before they carry you. If you didn’t have the results, they would test you at a fee. I remember it was USD 60 for the COVID-19 test before you get into an ambulance.”* (Nurse, HCW FGD, Harare).

Some healthcare workers believed that an increase in the number of home deliveries without a skilled birth attendant resulted in an increased number of largely unreported maternal and neonatal deaths.

> *“I don’t have statistics, but as I was on night duty, you would see that some people did not come for their regular reviews, some would come already complicated which if we had seen them on time, we could have prevented those complications."* (Nurse in charge, IDI, Bulawayo).

> *“I’m not sure in terms of statistics of women who could have been unfortunate to die during childbirth because of lack of access to health care, we don’t really have access to such information, but we know that happened.”* (CBO representative, KII, Bulawayo).

> *“I know plenty who needed maternity services but could not get them…in fact, I know someone who even died. They wanted to give birth…but they were supposed to get COVID-19 tests before accessing health services…that process was costly and trying to find money took long and she died before she could even be given medical attention.”* (Participant, household IDI, Harare).

Other studies reported similar findings of women and children struggling to access MCH services during the pandemic [10,68]. In Nyanga, Manicaland province, Nyashanu *et al* (2021) describe transport problems, roadblocks, shortage of medication and personal protective equipment, and lack of routine healthcare services affected routine health access such as antiretroviral therapy [41]. Chimhunya *et al* (2021) viewed strikes by healthcare workers at Sally Mugabe Hospital in Harare as an indirect consequence of COVID-19 which caused a decline in neonatal admissions [65].

#### (ii) Inter-personal and household relations

Another theme explored in our qualitative research was the impact of lockdown on interpersonal and domestic relationships against a backdrop of increasing stress and mental illness due to prolonged periods of home confinement and increasing levels of poverty and economic insecurity. Our findings indicated that gender-based violence (GBV) had increased.

> *"But with men, I would not say it’s domestic violence, I would have said I think men suffered emotional trauma because obviously when your woman is sitting at home, you are not providing as much as you used to, they would not be violent, but they would go through more of emotional trauma."* (Participant, community FGD, Harare).

> *“It could have been there before COVID-19 but with the lockdown restrictions the GBV got worse. So, those things are now like permanent scars in the community.”* (CBO representative, KII, Bulawayo).

> *“We also saw an upsurge of gender-based violence cases, some of them were reported through the normal channels, through the police and also health facilities but we also see an upsurge of some of these cases that were reported in the media”* (NGO representative, KII, Harare).

> *“Yes. You would notice that where we would expect that maybe the children, the girls are safe within the home, they were not even safe…We got reports of cases of rape by family members… Mobility restriction to survivors to access services then resulted in some cases of rape being reported late because people didn’t know."* (CBO representative, KII, Bulawayo).

These findings are supported by other studies. A cross-sectional online survey conducted in May 2020 among 507 Zimbabwean adults found signs and symptoms of generalised anxiety disorder present in almost half (40%) of the participants [71]. A qualitative study among rural women in Nyanga concluded that COVID-19 exacerbated domestic violence through the effects of financial stress, hunger, frustrations and spending more time together and that child marriages increased [72]. Others observed that as families spent more time in close contact while experiencing economic and job losses, women were exposed to more violence [73]. According to the Zimbabwe Gender Commission, domestic violence more than doubled [74] and some security personnel abused their authority by requesting favours from women in exchange for permission to sell their wares amidst lockdown restrictions [72].

Unfortunately, we were not able to obtain official data on reports of GBV to the police. To date, there is no official account of a rise in domestic violence in Zimbabwe. But even if such data showed a rise in the incidence of GBV, this would likely be an under-representation of the true picture because lockdown measures had created barriers to survivors of GBV making reports to the police or accessing services.

#### (iii) Social and economic well-being

Our primary research confirmed severe negative impacts on household income due to reduced economic activity and rising unemployment, a situation that was accentuated by the rising price of medicines, food, and other commodities [75]. One consequence was a reduction in both the number of meals and the quantity of food, disproportionately affecting those from the lowest socioeconomic strata, as well as women and children.

> *"On the issue of livelihoods, many people lost their jobs due to the closure of companies, most companies closed, even those that have since opened they are only taking a limited number. Most people were retrenched and came to just sit at home; this affected food security".* (Participant, Community FGD, Bulawayo).

> *“Then there was near starvation, we were not prepared for such a lockdown, it just emerged from nowhere, we were no longer able to travel, we were not able to buy enough food, some of us had no money, some of us were vendors we expected the day-to-day income"*, (FGD participant, Bulawayo).

> *“Livelihoods were affected in a way such that in families there was a shortage of food due to lack of money as some of the breadwinners became jobless…. Basic commodities were in short supply especially mealie meal and cooking oil.”* (Nurse, HCW FGD, Harare).

> *"Before the lockdown period, we would eat porridge in the morning, tea later, then have sadza [Zimbabwe staple food made from maize meal flour] for lunch. But because of lockdown we would eat once per day, or sometimes not find anything to eat at all.”* (Participant, household IDI, Bulawayo).

According to the World Bank, the pandemic added 1.3 million to the global number of people defined as extremely poor [76]. A cross-sectional online survey of 507 adults conducted at the beginning of the COVID-19 pandemic in May 2020 found that lockdown had increased food prices (95%) and decreased availability of nutritious foods (64%) [71]. The prevalence of wasting increased from 3.6% in 2019 to 4.5% in 2020 [20]. Other qualitative studies confirmed such negative impacts and noted a disproportionate negative impact on women whose caregiving roles had increased during lockdown [18,72].

#### (iv) Education and child wellbeing

School closure meant that many children received little to no formal school education during the COVID-19 lockdowns. Feelings of stress and anxiety about the lack of learning, especially for those in exam years, were expressed by many participants. Many informants also described how school and college/university closures widened existing educational inequalities between private versus public institutions and urban and rural settings because of the differential abilities to access teaching provided through online platforms and television or radio broadcasts [72].

> *"The government introduced in some schools online learning which does not apply to a rural child. And even if it’s urban, electricity, or issues of data, that’s a challenge as well. So, there are selected few students that can access school online depending on the background of the child…Then, there are radio programs that were scheduled and shared to all the schools…That was a good initiative, but again the issue of how many have radios, how many have electricity”* (CBO representative, KII, Bulawayo).

> *“The closing of schools resulted in a big divide between the poor and the rich, so the students who could afford extra lessons learnt and those who could not afford did not and the results were devastating”* (NGO representative, KII, Harare).

The long period of school closures, especially during the first lockdown, was also said to have increased smoking, use of alcohol and illicit drugs and sexual behaviour, as well as participation in so-called ‘gang behaviour’.

> *“Now we have a surge of drug abuse, school dropouts and the like. To be honest crimes have increased.”* (CBO representative, KII, Bulawayo).

> *“Since children were not going to school, they would spend time sitting at bridges, smoking marijuana, yet if they were in school, they could have been busy with their homework, but at that time they had no homework. The girls also were all over and ended up being impregnated.”* (Participant, household IDI, Harare).

> *“We work with children who are disadvantaged as a charity organization, but we noticed that during the lockdown, there were lots of unwanted pregnancies among teenagers and also alcohol abuse among the boys”* (Participant, household IDI, Harare).

We were unable to obtain routinely collected raw data on school attendance before and during the COVID-19 period. However, a 2022 Ministry of Primary and Secondary Education report provided some details on school enrolments, dropouts, transition, completion and examination pass rates over five years (2017-2021).[73] Gross Enrolment Ratio (GER) is a measure of participation in the education system and defined as the number of students enrolled at the beginning of the school year in specific levels of education, regardless of age, expressed as a percentage of the official eligible school-age population of a given level of education. Transition conveys information on the degree of access or transition from one cycle or level of education to a higher one, namely from Grade 7 to Form 1 and from Form 4 to Form 5. Completion rates indicate the proportion of persons completing a given level of education.

Primary education has two levels: infant (early child development (ECD) A/B and Grade 1/2) and junior (Grade 3-7) [77]. Transition into secondary school is after sitting for the Grade 7 national examination. The secondary education has also two levels, which are lower secondary (Form 1-4) and upper secondary (Form 5-6). Transition to upper secondary is only after attaining good grades at competitive national examinations. National ordinary level (‘O’ level) exams are completed after Form 4 and national advance (‘A’ level) exams are a requirement for university admission. While the total number of pupils enrolled in ECD and primary school have steadily increased from 2017-2021, the number of new entrants into Grade 1 decreased by more than 30,000 between 2020 and 2021. The number enrolled in higher secondary school also decreased by 14,941 (-14.7%) between 2020 and 2021. GER decreased across primary, lower secondary and higher secondary in 2020 and 2021 compared to previous years (Figure 4). Most other indicators also worsened in 2020 and 2021 compared to years 2017-2019 (which had seen a steady improvement year after year) including increased school dropouts, lower transition rates from Grade 7 to Form 1 and Form 4 to Form 6, lower completion rates across primary and secondary school and lower pass rates across all national exams. Worsening indicators seem to have affected boys and girls similarly.

**Figure 4:**
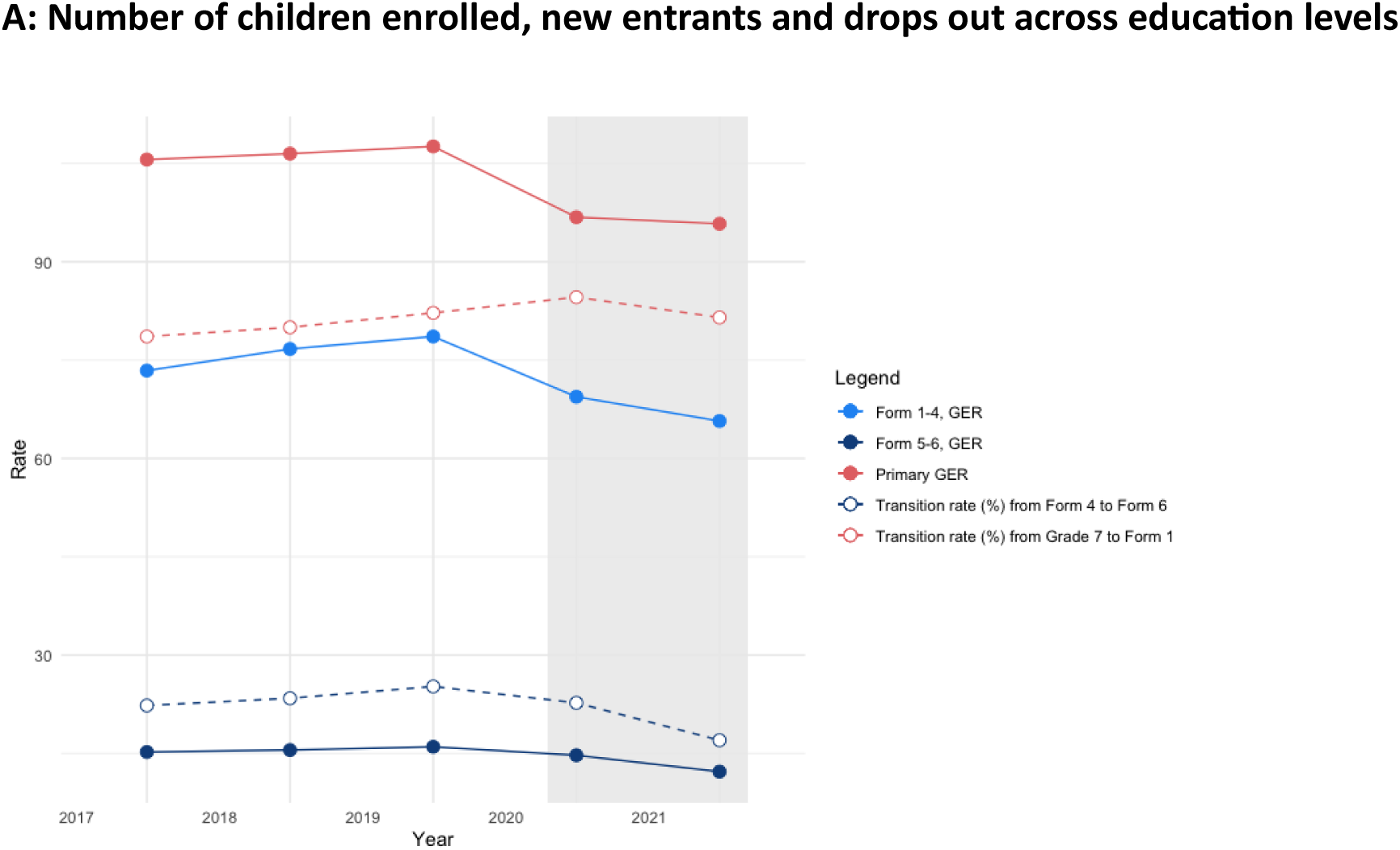
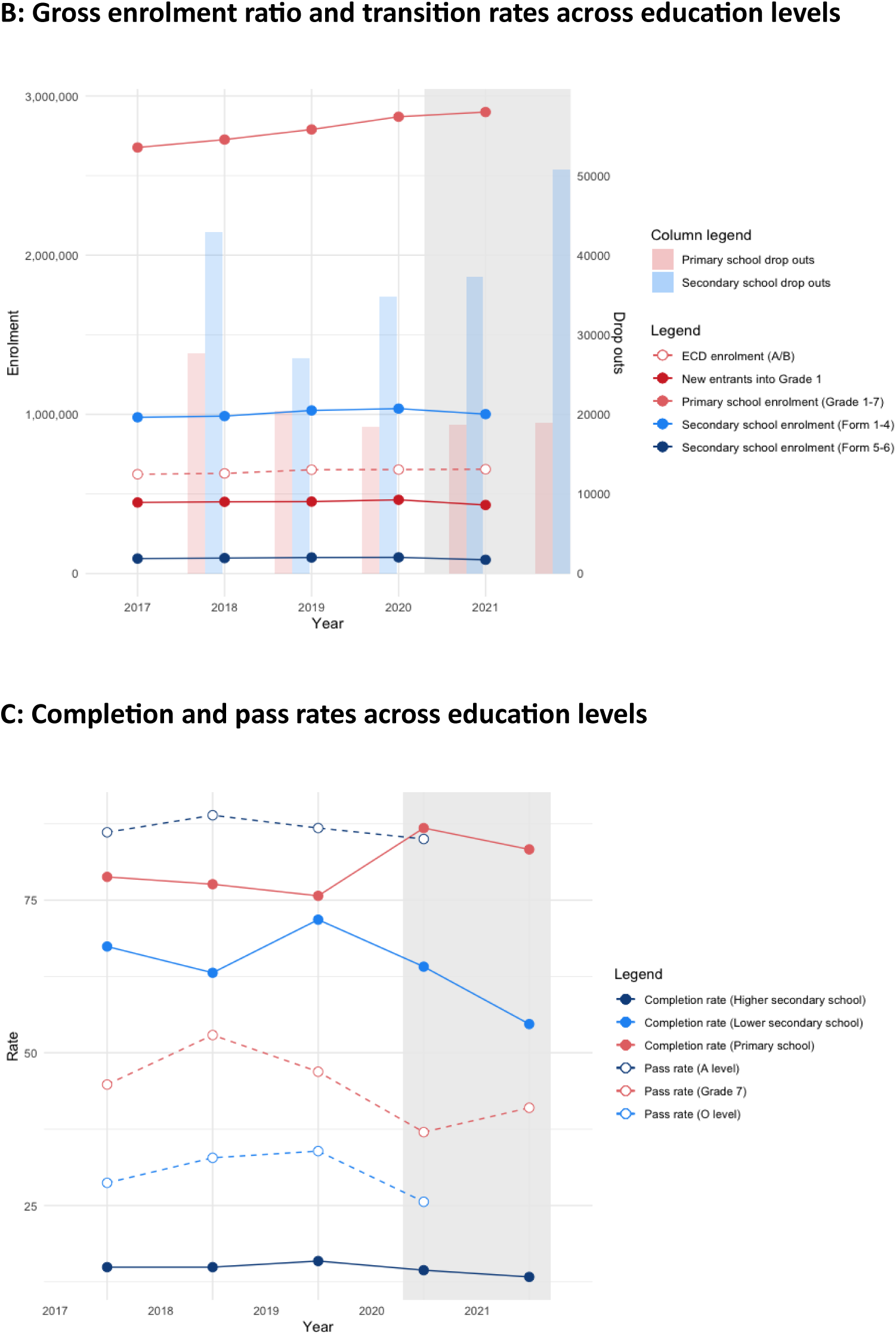
School indicators between 2017-2021.

## DISCUSSION

The COVID-19 pandemic occurred in Zimbabwe at a time when the country had experienced decades of economic decline and a chronic deterioration of its health system. Around seven out of every ten people were living below the international poverty line and health status indicators were among the poorest in Sub-Saharan Africa [16]. Furthermore, in the years immediately preceding the COVID-19 pandemic, the health system experienced intermittent industrial action by nurses and doctors, a series of natural disasters, and outbreaks of cholera, typhoid and measles [78]. Although the government increased its budgets for healthcare and unfroze healthcare worker posts to help cope with the pandemic [40], the fragility of the health system also made it likely that lockdown measures would only further reduce both the supply and demand for healthcare [79].

Given this background, Zimbabwe faced considerable challenges when COVID-19 emerged. As with all countries, policymakers had to contend with a lack of knowledge about the natural history, virulence and transmissibility of the virus [80]. The high rates of hospitalisation and mortality observed in China, Italy, and other countries as well as WHO’s declaration of a Public Health Emergency of International Concern on January 30^th^, 2020, led many countries to implement stringent lockdown measures before they experienced any upsurge in COVID-19 cases. Underpinning these decisions was the belief amongst public health specialists and policymakers that any harms produced through the imposition of lockdown measures would be less than the harms produced by the virus itself.

Early estimates of the infection fatality rate were such that the enforcement of lockdown, including the cessation of important social and economic activities, was felt to be justified. However, from the middle of 2020, differences in the incidence of cases and deaths were being observed across the world [62]. The relatively low incidence of cases and deaths in parts of Africa was especially notable [62] leading to several hypotheses as to what might be the cause. Among them was the low prevalence of risk factors such as obesity, low infection rate, effective mitigation measures, youthful age structure, favourable warm weather, and preexisting immunity from previous coronavirus infection. In Zimbabwe, by the end of 2020, only 13,867 cases and 363 deaths had been officially recorded and by the end of 2021, the number of deaths attributed to COVID-19 was only 5,004 [81]. While there would have been many unrecorded deaths due to COVID-19, there was no period when hospitals were overrun by COVID-19 patients or when morgues struggled to cope with a sudden rise in deaths as experienced by many other countries. Furthermore, seroprevalence studies indicate that substantial transmission had occurred by the end of 2020 [59], indicating that the relatively low mortality rates were possibly not due to effective prevention of transmission.

The importance of reliable, timely and context-specific data to manage infectious diseases outbreaks and epidemics cannot be understated. In Zimbabwe, the gaps in the health information system meant that a timely, accurate and detailed account of the epidemic and its direct impacts were unavailable. Throughout the pandemic, policymakers and public health specialists had to make educated guesses as to how the epidemic was evolving and what impact it was having. The not-so-perfect health information systems in the country also meant that policymakers and public health specialists could not adequately assess the harmful and sometimes devastating impact of the lockdown measures. Even now, estimates of the size and the causes of the increase in all-cause mortality since the pandemic are unclear.

A key question emerging from this research is whether Zimbabwe adopted the right set of measures to effectively manage both the direct and indirect threats to health that were posed by COVID-19, including the harms associated with lockdown. Here one must be wary of the benefit of hindsight and the lack of reliable and timely data available to policymakers and public health experts at the time. However, given the pre-existing and high levels of poverty and food insecurity before the pandemic [17], the economic stagnation, reduced household income and rising prices for basic commodities caused by the lockdowns were inevitably going to be impactful. The inability of many households to generate an adequate income during lockdown had drastic and detrimental effects on access to food and basic amenities and was compounded by mounting loneliness, distress, and mental illness, fuelling a degradation of social relations within households and an increase in interpersonal violence and antisocial behaviour. Although the government created a support fund for poorer households, the budget was inadequate, and many eligible households did not receive the promised support. Studies from other LMICs have shown similar findings [6,8,82] and many countries are currently still experiencing the long-term consequences of the economic contraction and increased levels of indebtedness that occurred in 2020 and 2021 [83].

Similarly, a negative impact on an already fragile, under-resourced and under-utilised health system was inevitable as were the significant and long-term negative effects on students and their families due to the prolonged closure of education facilities. The disproportionate impact on children is of particular importance. While COVID-19 was considerably much less harmful to children, the converse is true for the effects of the lockdown measures, including the cessation or reduction of essential child healthcare services; the heightened vulnerability of infants and young children to reductions in food intake; and the cessation of school. Thus, many children were deprived of essential care and services as a result of measures that mostly benefited adults.

A further important point is that for large segments of the population, lockdown measures were not feasible. Many informants explained how safe and adequate social distancing was impractical in high-density areas and informal settlements. If true, for many households, prolonged quarantine and social isolation were arguably unjustified, especially considering the harmful effects of lockdown measures described above. Moreover, lockdown measures of the kind experienced with COVID-19 involve extraordinary curtailments on civil liberties that should only be implemented in extreme and valid circumstances. The coercive powers claimed by governments across the world in the name of public safety may not only be abused during a pandemic but also used to establish permanent systems and mechanisms of surveillance and social control. Indeed, lockdown measures were used in Zimbabwe to curtail political protest, and there were also accounts of security sector actors abusing their power to commit corruption and sexual harassment to the population.

Firm conclusions about the full impact of COVID-19 and its associated communicable disease control measures are not possible due to the lack of reliable and complete data. Crucially, any future strengthening of the health information system must include a focus not just on the future ability to conduct infectious diseases surveillance and control, but also on the ability to avoid or mitigate negative social, economic, and educational impacts. Nonetheless, plenty of data and evidence exist to suggest that the indirect impacts of COVID-19 were at least as harmful if not greater than its direct impacts especially so for women and children.

## Data Availability

Data will be made available free access and a DOI will be provided once available. Data is deposited at the London School and Hygiene and Tropical Medicine (LSHTM) data campass

## Authors’ contributions

TT, JD, KK, and DM conceptualized the study. IO, TB, TT, MM, and KM cleaned and curated the data. TT, IO, RSC, AOY, and NO with support from KD, KK, JD, and DM analysed the data. DM, KW, and RAF acquired the funds for the study. SS, LM, CM, and SM supported by RSC, AYO, NO, AM, and TT performed the qualitative data collection. Study coordination and supervision of field teams were done by RSC, TT, SN, ACM and TC. TT wrote the first draft of the manuscript with feedback from JD, KK, and DM. All authors provided feedback on the manuscript and read and approved the final version of the manuscript.

## Acknowledgements

The study would like to acknowledge the support we got from the Zimbabwe research authorities, government departments and local authorities. In addition, the study also acknowledges Professor John Metcalfe, Thokozile Masha, Jane Mususa, and the OPHID and BRTI team led by Dr Shungu Munyati, Thomas Godfrey-Faussett who helped with valuable reference materials on education and most importantly the participants who spared their time to share this valuable experience. AYO, OD, is a member of and acknowledges support from, the Pan-African Network for Rapid Research, Response, Relief and Preparedness for Infectious Disease Epidemics, funded by the European and Developing Countries Clinical Trials Partnership, under the EU Horizon 2020 Framework Programme for Research and Innovation.

